# COVID-19 Vaccination Booster Uptake Is Related to End of Pandemic Perception: Population-Based Survey in Four Provinces in Indonesia

**DOI:** 10.1101/2024.03.05.24303828

**Authors:** Ahmad Watsiq Maula, Citra Indriani, Risalia Reni Arisanti, Vincentius Arca Testamenti, Catharina Yekti Praptiningsih, Rebecca Merril, Ansariadi, Tri Yunis Miko Wahyono, Atik Choirul Hidajah, Riris Andono Ahmad

## Abstract

Global COVID-19 booster vaccination uptake has been low, particularly in low-middle-income countries (LMICs). However, studies on the determinants of COVID-19 booster vaccination remain limited, especially in LMIC settings. This study aims to describe the determinants of COVID-19 booster vaccination uptake in an LMIC context. We analyzed data from a cross-sectional survey that was conducted in September 2022 in four provinces in Indonesia. Participants (n=2,223) were recruited using multiple-stage cluster sampling. Multivariable logistic regression analysis was used to identify factors associated with booster vaccination status. The proportion of COVID-19 booster vaccination among fully vaccinated adults was 29.5%, while fear of transmission (12.4%) and perceived risk of getting a COVID-19 infection (33.5%) were low. Multivariable analysis showed that people living on Java Island (aOR: 2.45, 95% CI: 1.87-3.24), living in the urban area (aOR: 2.04, 95% CI: 1.60-2.61), being an employee in the formal sector (aOR: 3.99, 95% CI: 1.93 - 8.58), experiencing a side effect from previous vaccination (aOR: 1.71, 95% CI: 1.40-2.09), having a history of SARS-COV2 infection (aOR: 2.10, 95%CI: 1.27-3.50), having perception on the upcoming new wave of COVID-19 (aOR: 1.37, 95% CI: 1.07 -1.76), and believing the pandemic has not ended (aOR: 1.29, 95% CI: 1.01, 1.64) were associated with booster shot uptake. Low educational level (aOR: 0.6, 95% CI: 0.39-0.93) inhibited booster vaccination uptake. Current booster dose coverage was considerably lower than the primary vaccination dosage. The low booster vaccination uptake in four provinces was associated with a belief the pandemic has no longer, which might hinder the catch up of wide-population target coverage and COVID-19 control for reduced disease severity and hospitalization. Thus, efforts need to be prioritized in reaching the COVID-19 vulnerable population, which includes elderly and those with comorbidities.

## Introduction

Implementing the COVID-19 vaccination program was one of the most effective interventions to control the ongoing COVID-19 pandemic (Shakeel et al., 2022). However, the immunity conferred by the initial vaccination waned significantly after 8 and 11 months of the first dosage. As a result, booster vaccination is required to prevent severe illness and mortality (Hernandez-Suarez and Murillo-Zamora, 2022). In addition, studies showed that COVID-19 boosters provided additional protection against Delta and Omicron variants of SARS-CoV-2, reducing the risk of emergency visits and hospitalization (Thompson, 2022; Tseng et al., 2022).

Increasing uptake of a booster vaccination is currently recommended worldwide to prepare for the potential resurgence of COVID-19 due to a new circulating variant to prevent death and reduce transmission among those at most significant risk (Betsch et al., 2022; Lu et al., 2023; Machado et al., 2022; Moola et al., n.d.). However, despite high primary vaccination campaign coverage, global data reveals a slow and unequal rollout of booster doses, particularly in low-income and middle-income countries (Schellekens, 2022).

Vaccine hesitancy was the key factor for the slow uptake of COVID-19 booster vaccines in 23 countries in Europe and Asia (Lazarus et al., 2023, 2021; Moola et al., n.d.; Sallam, 2021). Many factors underlying the discouragement from receiving vaccines include lack of trust in government, the prospect of job loss, education level, lack of awareness, perceived immunity, risk perception, concern and uncertainty on safety and effectiveness (Abouzid et al., 2022; Bennett et al., 2022; Sinclair, 2023).

In Indonesia, the Ministry of Health recommended COVID-19 vaccine booster doses for health workers on July 23, 2021, and expanded the eligibility to all individuals aged 18 or more by January 12, 2022. By May 2022, the primary vaccine dose coverage was 70%. However, the booster dose coverage was only 24.8%. This lower coverage was despite two studies in Indonesia that found the acceptance of booster dosage was high at 56.3% and 93.9% (Harapan et al., 2022; Wirawan et al., 2022). Understanding the factors contributing to booster shot hesitancy is essential to help policymakers optimize vaccination strategies for COVID-19 control. This study in four provinces in Indonesia aimed to assess COVID-19 booster vaccination coverage and identify factors that cause adults with complete primary series vaccination to hesitate receiving booster dosage.

## Methods

### Study participants

The data used in this study was part of a population-based household cross-sectional prevalence study conducted in Indonesia in September 2022 that will be reported elsewhere. The cross-sectional study was a national-level survey designed to assess the prevalence of SARS-CoV-2 infection among government targeted populations in four provinces, Banten, West Java, East Java, and South Sulawesi. Multistage cluster sampling was used to enroll 3,626 participants from 1,865 households in 144 clusters distributed in 54 districts and cities, with 79% of participants living in rural areas. A cluster was defined as a hamlet comprising 50 to 90 households on average. All household members aged five years and above were invited to the study.

### Study sites

Data collection was conducted in four provinces in Indonesia; Banten Province, West Java Province, East Java Province, and South Sulawesi Province, with a total number of inhabitants corresponding to 41.6% of the national population (Figure 1). Additionally, these provinces are listed in the top ten contributors of national COVID-19 cases in 2020–2022 in Indonesia, accounting for 1.812951 (35.2%) cases [https://data.covid19.go.id/public/api/prov.json]. Indonesia is an archipelago comprising five main islands. Java Island is the geographic and economic center of Indonesia, where more than half of Indonesia’s population lives.

**Figure 1.**
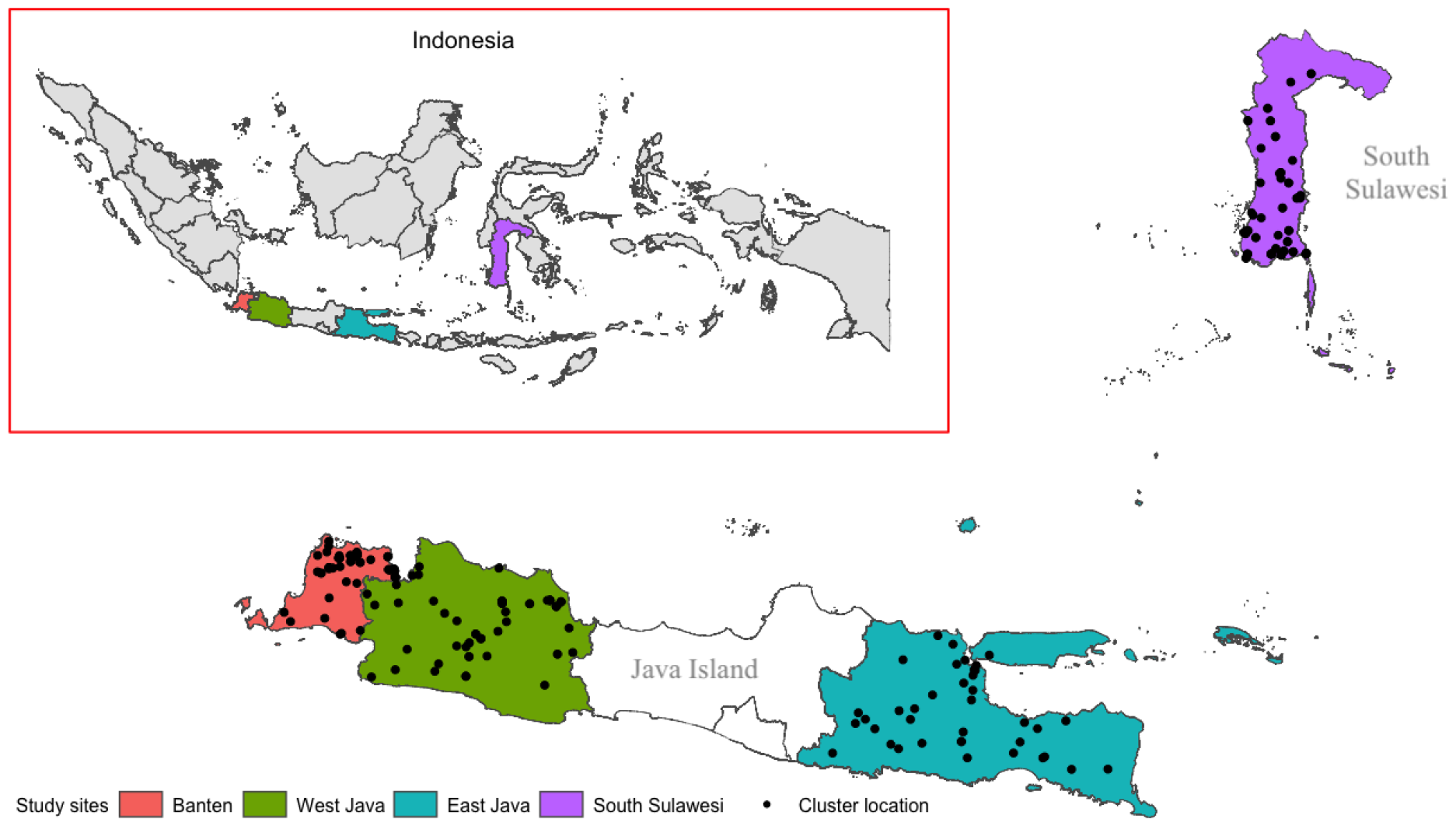
The study site and cluster distribution for a cross-sectional survey in four provinces in Indonesia. Three provinces are in Java Island and a province located in the Sulawesi Island.

### Sample size

Subset samples of participants aged 18 years and older who were eligible for the booster of COVID-19 vaccination (Figure 2) was selected from study database. There were 41 participants excluded from the analysis due to ineligibility for a booster vaccine (34), i.e., they had received the second dose less than three months prior and had no information on the date of the second dosage vaccination (7). Thus, we included data from 2.223 participants in the analysis.

**Figure 2.**
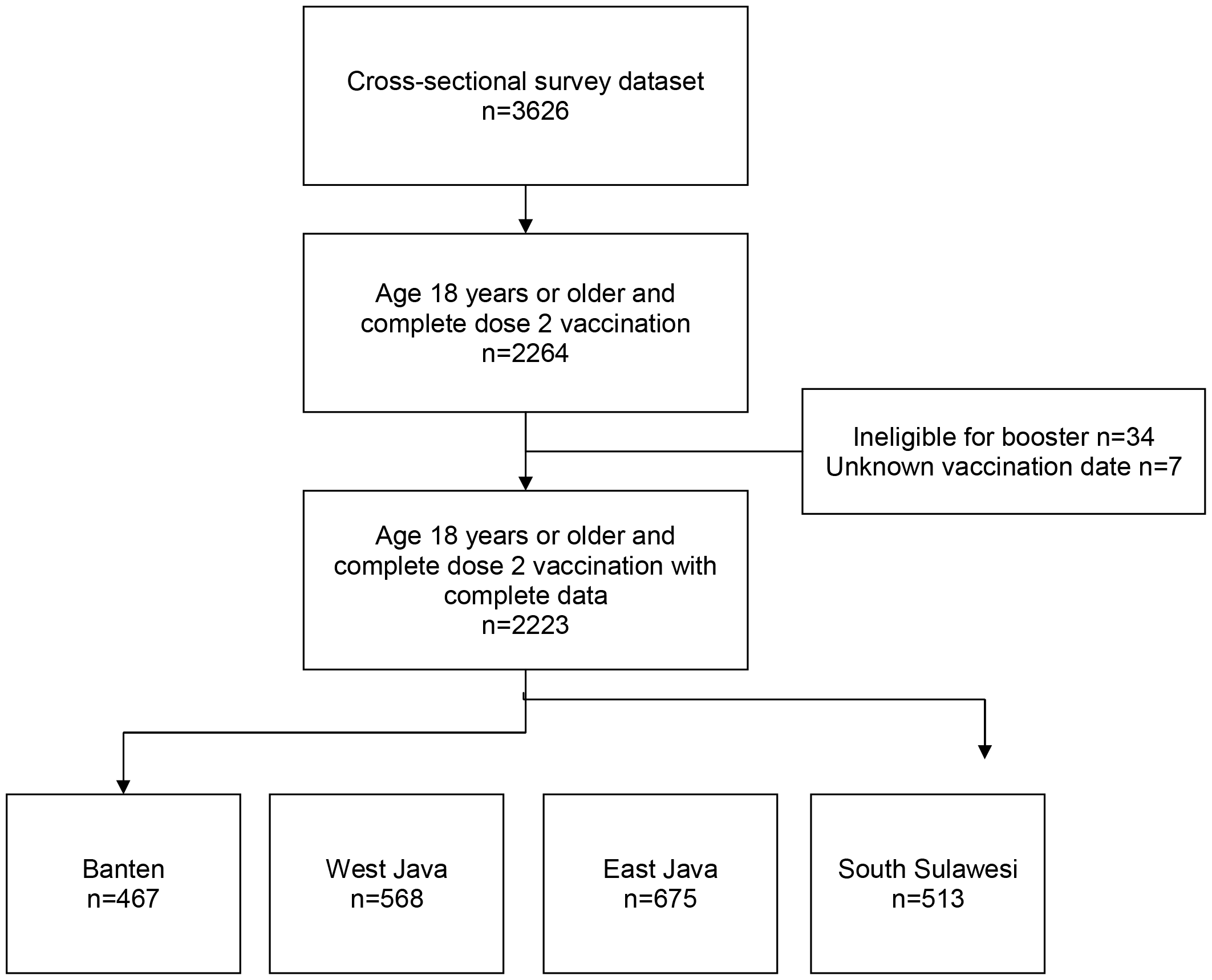
Flow chart describing participants’ sample size included in the study from the original data set. Sample size varies depending on the data collection for each sub-area.

**Figure 3.**
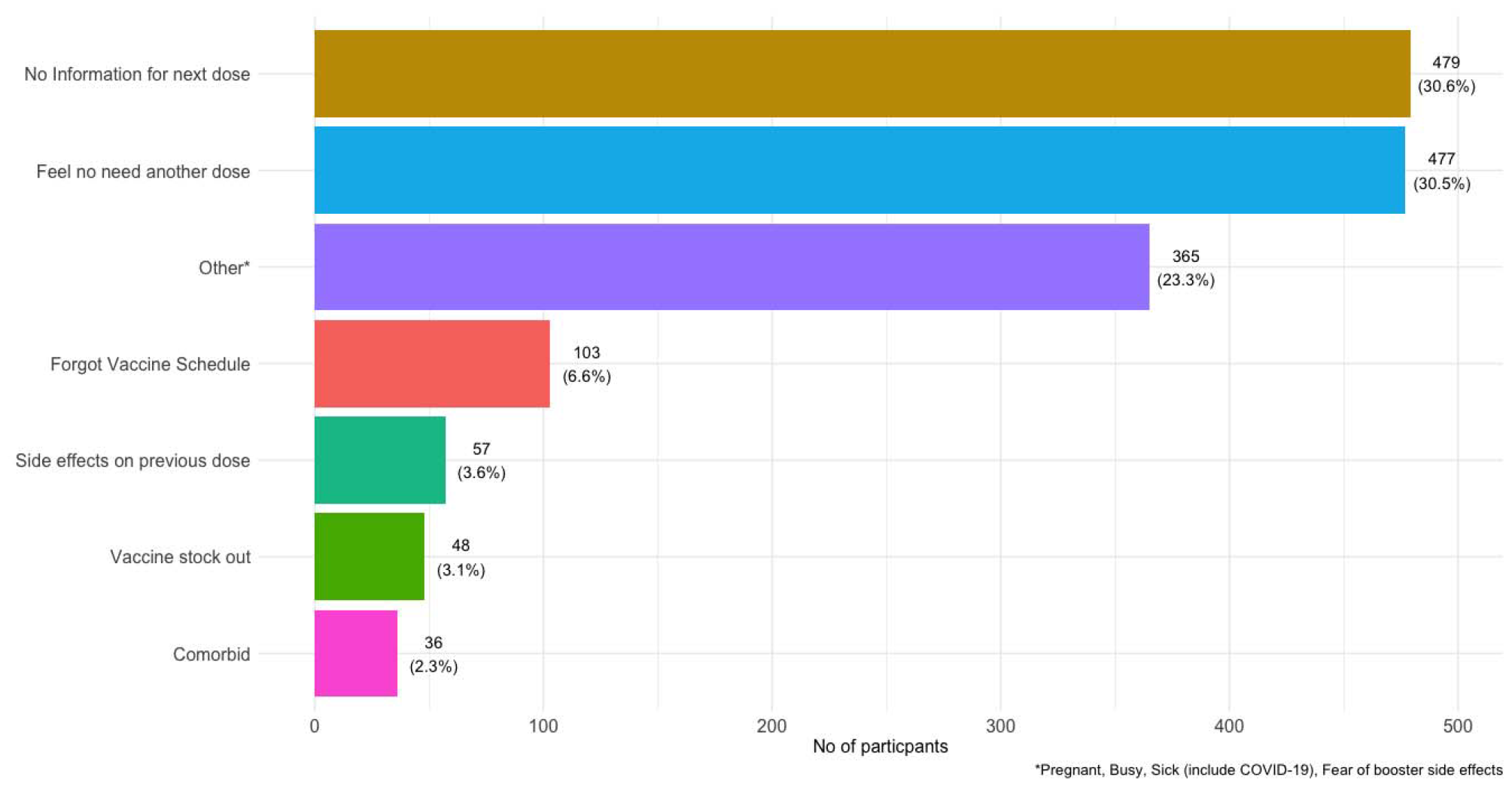
Reason for not getting booster dosage shot among adults complete the primary vaccine series.

#### Data collection and management

Trained enumerators used a structured paper-based questionnaire to collect demographic data, existing comorbidities, booster vaccination status, perception towards individual risk of being infected SARS-CoV-2, perception of an upcoming new COVID-19 wave, and the end of the pandemic through face-to-face interviews. Supervisors validated the collected data daily and entered it into an electronic database using the KOBO Toolbox, available at https://www.kobotoolbox.org/. The data manager checked the data consistency and completeness on a daily basis.

### Statistical analysis

We compare the vaccination booster status on demographic characteristics (age, gender, education, occupation, household income, living area), existing comorbidity, and perception towards pandemic transmission situation and risk of getting COVID-19 infection. We used multiple logistic regression to calculate the odds ratio (OR) and adjusted OR (aOR) for other covariates and assessing the interaction. The Wald Chi Squared Test with p-value <0.05 considered as statistically significant for association and the Likelihood-Ratio Test with p-value <0.01 indicated a significant interaction. In addition, we performed the variance inflation factor (VIF) measurement to detect multicollinearity between variables in the logistic regression models. The cut-off value for multicollinearity used in this study was 5. We used R software version 4.2.2 (R Foundation for Statistical Computing, Vienna, Austria) for data analysis.

### Research ethics

The study protocol ethical approval was received from the Medical and Health Research Ethics Committee of the Faculty of Medicine, Public Health, and Nursing Universitas Gadjah Mada (No. KE/FK/1139/EC). It was acknowledged by the Indonesian National Research and Innovation Agency and has been determined to fall under the WHO Unity umbrella determination at U.S. Centers for Disease Control and Prevention. Written informed consent was obtained from each participant before collecting data.

## Results

### Baseline characteristics of participants

The study population consisted of 66.0% (n=1,467) females and 34.0% (756) males, with 53.4% (1,188) seen in the age group 18 – 44 years old. Most of the study population were living in rural areas (79.0%) and had either elementary school (36.4%) or secondary school (47.9%) education. Most participants worked in the informal sector (43.9%) and as a housewife (37.1%). More than half participants (59.2%) had no reported comorbidity. The complete characteristics of the study participants are presented in Table 1. Generally, booster vaccination coverage was 29.5% but varied between provinces, with higher coverage in provinces on Java Island (25.1% – 37.7%) and lower South Sulawesi in Sulawesi Island (17.9%).

**Table 1.** Descriptive characteristics of the participants included in the study (n=2.223)

| Characteristic | Java Island |  |  | Outside Java Island | Total |
| --- | --- | --- | --- | --- | --- |
|  | West Java | Banten | East Java | South |  |

|  |  |  |  |  | Sulawesi |  |
| --- | --- | --- | --- | --- | --- | --- |
|  |  | n=568 | n=467 | n=675 | n=513 | n=2.223 |
|  |  | n(%) | n(%) | n(%) | n(%) | n(%) |
| Booster status |  |  |  |  |  |  |
|  | Primary vaccination | 354 (62.3) | 350 (74.9) | 443 (65.6) | 421 (82.1) | 1568 (70.5) |
|  | Booster | 214 (37.7) | 117 (25.1) | 232 (34.4) | 92 (17.9) | 655 (29.5) |
| Age group |  |  |  |  |  |  |
|  | 18-44 years old | 300 (52.8) | 284 (60.8) | 324 (48.0) | 280 (54.6) | 1188 (53.4) |
|  | 45-59 years old | 179 (31.5) | 128 (27.4) | 237 (35.1) | 168 (32.7) | 712 (32.0) |
|  | >=60 years old | 89 (15.7) | 55 (11.8) | 114 (16.9) | 65 (12.7) | 323 (14.5) |
| Residential area |  |  |  |  |  |  |
|  | Urban | 117 (20.6) | 154 (33.0) | 64 (9.5) | 132 (25.7) | 467 (21.0) |
|  | Rural | 451 (79.4) | 313 (67.0) | 611 (90.5) | 381 (74.3) | 1756 (79.0) |
| Sex |  |  |  |  |  |  |
|  | Male | 178 (31.3) | 156 (33.4) | 248 (36.7) | 174 (33.9) | 756 (34.0) |
|  | Female | 390 (68.7) | 311 (66.6) | 427 (63.3) | 339 (66.1) | 1467 (66.0) |
| Household Income (IDR) |  |  |  |  |  |  |
|  | < 1.000.000 | 116 (20.4) | 99 (21.2) | 101 (15.0) | 159 (31.0) | 475 (21.4) |
|  | 1.000.000-4.999.999 | 307 (54.0) | 215 (46.0) | 433 (64.1) | 255 (49.7) | 1210 (54.4) |
|  | 5.000.000-9.999.999 | 105 (18.5) | 91 (19.5) | 103 (15.3) | 73 (14.2) | 372 (16.7) |
|  | >= 10.000.000 | 40 (7.0) | 62 (13.3) | 38 (5.6) | 26 (5.1) | 166 (7.5) |
| Education |  |  |  |  |  |  |
|  | No formal education | 38 (6.7) | 18 (3.9) | 38 (5.6) | 51 (9.9) | 145 (6.5) |
|  | Elementary school | 261 (46.0) | 160 (34.3) | 220 (32.6) | 169 (32.9) | 810 (36.4) |
|  | Secondary school | 221 (38.9) | 245 (52.5) | 370 (54.8) | 229 (44.6) | 1065 (47.9) |
|  | Higher education | 48 (8.5) | 44 (9.4) | 47 (7.0) | 64 (12.5) | 203 (9.1) |
| Occupation |  |  |  |  |  |  |
|  | Unemployed | 60 (10.6) | 76 (16.3) | 106 (15.7) | 69 (13.5) | 311 (14.0) |
|  | Student | 9 (1.6) | 16 (3.4) | 15 (2.2) | 17 (3.3) | 57 (2.6) |
|  | Housewives | 261 (46.0) | 172 (36.8) | 195 (28.9) | 196 (38.2) | 824 (37.1) |
|  | Civil servant/army/police/health worker | 8 (1.4) | 9 (1.9) | 16 (2.4) | 21 (4.1) | 54 (2.4) |
|  | Entrepreneur, farmer, fisherman, daily worker and other | 230 (40.5) | 194 (41.5) | 343 (50.8) | 210 (40.9) | 977 (43.9) |
| Self-reported existing comorbidities |  |  |  |  |  |  |
|  | Yes | 239 (42.1) | 191 (40.9) | 262 (38.8) | 215 (41.9) | 907 (40.8) |
|  | No | 329 (57.9) | 276 (59.1) | 413 (61.2) | 298 (58.1) | 1316 (59.2) |

**Table 2.** Individual perception toward COVID-19 transmission, upcoming new wave, and the end of the pandemic among 18 years old and older who complete the second dose of vaccination in four provinces.

| Individual Perception of the COVID-19 Pandemic |  | West Java | Banten | East Java | South Sulawesi | Total |
| --- | --- | --- | --- | --- | --- | --- |
|  |  | n (%) | n (%) | n (%) | n (%) | N (%) |
| Fears of COVID-19 transmission |  |  |  |  |  |  |
|  | Fears | 93 (16.4) | 48 (10.3) | 82 (12.1) | 52 (10.1) | 275 (12.4) |
|  | Neutral | 163 (28.7) | 132 (28.3) | 187 (27.7) | 102 (19.9) | 584 (26.3) |
|  | No fears | 312 (54.9) | 287 (61.5) | 406 (60.1) | 359 (70.0) | 1364 (61.4) |
| Fears of getting infected by SARS-Cov2 |  |  |  |  |  |  |
|  | At risk | 223 (39.3) | 191 (40.9) | 193 (28.6) | 137 (26.7) | 744 (33.5) |
|  | Not at risk | 250 (44.0) | 212 (45.4) | 380 (56.3) | 253 (49.3) | 1095 (49.3) |
|  | Do not know | 72 (12.7) | 42 (9.0) | 67 (9.9) | 103 (20.1) | 284 (12.8) |
|  | Do not care | 23 (4.0) | 22 (4.7) | 35 (5.2) | 20 (3.9) | 100 (4.5) |
| Perception of COVID-19 new wave will come |  |  |  |  |  |  |
|  | Will come | 173 (30.5) | 124 (26.6) | 131 (19.4) | 82 (16.0) | 510 (22.9) |
|  | Will not come | 255 (44.9) | 217 (46.5) | 414 (61.3) | 246 (48.0) | 1132 (50.9) |
|  | Do not know | 140 (24.6) | 126 (27.0) | 130 (19.3) | 185 (36.1) | 581 (26.1) |
| Perception that the COVID-19 pandemic has ended |  |  |  |  |  |  |
|  | Agree | 259 (45.6) | 233 (49.9) | 359 (53.2) | 246 (48.0) | 1097 (49.3) |
|  | Disagree | 187 (32.9) | 96 (20.6) | 177 (26.2) | 93 (18.1) | 553 (24.9) |
|  | Do not know | 122 (21.5) | 138 (29.6) | 139 (20.6) | 174 (33.9) | 573 (25.8) |

### Individual Perception of the COVID-19 Pandemic

The proportion of those who feared COVID-19 transmission ranged from 10.1% to 16.4%, with the lowest level of fear observed in South Sulawesi and the highest in West Java. One-third of participants (33.5%) acknowledged being at risk of getting SARS-CoV-2 virus infection, while nearly half (49.3%) reported no longer being at risk. Half of the participants (50.9%) believed that Indonesia would not experience a new wave of COVID-19, and 45.6 - 53.2% had the perception that the COVID-19 pandemic had ended.

### Booster status among participants and hesitancy factors for booster uptake

Table 3 reports the adjusted odds ratio (aOR) from the logistic model predicting booster hesitancy among those who complete the primary dose series. We found that having a low education level was associated with the lower intention to have booster vaccination significantly (aOR: 0.6, 95% CI: 0.39-0.93) compared to high education level. Our result also showed that those living in Java (aOR: 2.45, 95% CI: 1.87-3.24) compared to South Sulawesi, living in an urban area (aOR: 2.04, 95% CI: 1.60-2.61) compared to rural area; being an employee in the formal sector (aOR: 3.99, 95% CI: 1.93 - 8.58) compare to the informal sector were strongly exhibited the booster vaccination shot compared than who have primary vaccination; Experiencing a side effect from previous vaccination (aOR: 1.71, 95% CI: 1.40-2.09); having a history of SARS-COV-2 infection (aOR: 2.10, 95%CI:1.27-3.50); having perception on the upcoming new COVID-19 wave (aOR: 1.37, 95% CI: 1.07 -1.76); and believing that the pandemic has not ended (aOR: 1.29, 95% CI: 1.01, 1.64) were associated with booster vaccination status.

**Table 3.** Vaccination booster status based on sociodemographic and associated factors.

| Variables<br>n(%) | Not Booster<br>1568 (70.5) | Booster<br>655 (29.5) | Total<br>2223 (100) | Crude OR<br>(95% CI) | Adjusted<br>OR (95% CI) |
| --- | --- | --- | --- | --- | --- |
| Island |  |  |  |  |  |
| Java | 1147 (73.2%) | 563 (86.0%) | 1710 (76.9%) | 2.25 *** (1.76, 2.89) | 2.45 *** (1.87, 3.24) |
| Outside Java | 421 (26.8%) | 92 (14.0%) | 513 (23.1%) | Ref |  |
| Residential area |  |  |  |  |  |
| Urban | 248 (15.8%) | 219 (33.4%) | 467 (21.0%) | 2.67 *** (2.16, 3.30) | 2.04 *** (1.60, 2.61) |
| Rural | 1320 (84.2%) | 436 (66.6%) | 1756 (79.0%) | Ref |  |
| Age (years) |  |  |  |  |  |
| >=60 | 225 (14.3%) | 98 (15.0%) | 323 (14.5%) | 1.12 (0.86 – 1.47) |  |
| 45-59 | 487 (31.1%) | 225 (34.4%) | 712 (32.0%) | 1.19 (0.97 – 1.46) |  |
| 18-44 | 856 (54.6%) | 332 (50.7%) | 1188 (53.4%) | Ref |  |
| Gender |  |  |  |  |  |
| Female | 1031 (65.8%) | 436 (66.6%) | 1467 (66.0%) | 1.04 (0.86, 1.26) |  |
| Male | 537 (34.2%) | 219 (33.4%) | 756 (34.0%) | Ref |  |
| Education |  |  |  |  |  |
| Elementary school | 642 (40.9%) | 168 (25.6%) | 810 (36.4%) | 0.76 (0.51, 1.16) | 0.60 * (0.39, 0.93) |
| Secondary school | 730 (46.6%) | 335 (51.1%) | 1065 (47.9%) | 1.34 (0.91, 2.01) | 0.80 (0.53, 1.24) |
| Higher education | 88 (5.6%) | 115 (17.6%) | 203 (9.1%) | 3.81 *** (2.41, 6.13) | 1.50 (0.88, 2.58) |
| No formal education | 108 (6.9%) | 37 (5.6%) | 145 (6.5%) | Ref |  |
| Household Income per month (IDR) |  |  |  |  |  |
| 1.000.000- 4.999.999 | 895 (57.1%) | 315 (48.1%) | 1210 (54.4%) | 1.14 (0.89, 1.47) | 0.89 (0.68, 1.16) |
| 5.000.000-9.999.999 | 225 (14.3%) | 147 (22.4%) | 372 (16.7%) | 2.12 *** (1.58, 2.85) | 1.22 (0.87, 1.69) |
| >=10.000.000 | 85 (5.4%) | 81 (12.4%) | 166 (7.5%) | 3.09 *** (2.13, 4.48) | 1.35 (0.88, 2.06) |
| <1.000.000 | 363 (23.2%) | 112 (17.1%) | 475 (21.4%) | Ref |  |
| **Occupation** |  |  |  |  |  |
| Student | 39 (2.5%) | 18 (2.7%) | 57 (2.6%) | 1.21 (0.65, 2.20) | 0.95 (0.48, 1.81) |
| Housewives | 599 (38.2%) | 225 (34.4%) | 824 (37.1%) | 0.98 (0.74, 1.32) | 1.11 (0.82, 1.53) |
| Civil servant/army/<br>police/health worker | 15 (1.0%) | 39 (6.0%) | 54 (2.4%) | 6.80 *** (3.64, 13.33) | 3.99 *** (1.93, 8.58) |
| Entrepreneur, farmer,<br>fisherman, daily<br>worker and other | 690 (44.0%) | 287 (43.8%) | 977 (43.9%) | 1.09 (0.82, 1.45) | 1.01 (0.74, 1.37) |
| Unemployed | 225 (14.3%) | 86 (13.1%) | 311 (14.0%) | Ref |  |
| **Self-reported existing comorbidities** |  |  |  |  |  |
| Yes | 639 (40.8%) | 268 (40.9%) | 907 (40.8%) | 1.01 (0.84, 1.21) |  |
| No | 929 (59.2%) | 387 (59.1%) | 1316 (59.2%) | Ref |  |
| **Vaccine side effects** |  |  |  |  |  |
| Yes | 716 (45.7%) | 405 (61.8%) | 1121 (50.4%) | 1.93*** (1.60, 2.32) | 1.71 *** (1.40, 2.09) |
| No | 852 (54.3%) | 250 (38.2%) | 1102 (49.6%) | Ref |  |
| **Previous COVID-19 Infection** |  |  |  |  |  |
| Yes | 31 (2.0%) | 55 (8.4%) | 86 (3.9%) | 4.54 *** (2.92, 7.20) | 2.10 ** (1.27, 3.50) |
| No | 1537 (98.0%) | 600 (91.6%) | 2137 (96.1%) | Ref |  |
| **Fears of COVID-19 transmission** |  |  |  |  |  |
| Fears | 191 (12.2%) | 84 (12.8%) | 275 (12.4%) | 1.05 (0.79, 1.38) |  |
| Neutral | 417 (26.6%) | 167 (25.5%) | 584 (26.3%) | 0.95 (0.77, 1.18) |  |
| No fears | 960 (61.2%) | 404 (61.7%) | 1364 (61.4%) | Ref |  |
| **Fears of getting infected with SARS-COV-2** |  |  |  |  |  |
| At risk | 485 (30.9%) | 259 (39.5%) | 744 (33.5%) | 1.30 * (1.06, 1.59) | 0.91 (0.72, 1.13) |
| Do not know | 230 (14.7%) | 54 (8.2%) | 284 (12.8%) | 0.57 *** (0.41, 0.78) | 0.76 (0.53, 1.08) |
| Do not care | 77 (4.9%) | 23 (3.5%) | 100 (4.5%) | 0.73 (0.44, 1.16) | 0.86 (0.51, 1.41) |
| Not at risk | 776 (49.5%) | 319 (48.7%) | 1095 (49.3%) | Ref |  |
| **Perception of COVID-19 new wave will come** |  |  |  |  |  |
| Yes | 296 (18.9%) | 214 (32.7%) | 510 (22.9%) | 1.83 *** (1.47, 2.28) | 1.37 * (1.07, 1.76) |
| Do not know | 460 (29.3%) | 121 (18.5%) | 581 (26.1%) | 0.67 *** (0.52, 0.85) | 0.85 (0.64, 1.11) |
| No | 812 (51.8%) | 320 (48.9%) | 1132 (50.9%) | Ref |  |
| **Perception that the COVID-19 pandemic has ended** |  |  |  |  |  |
| Disagree | 431 (27.5%) | 142 (21.7%) | 573 (25.8%) | 1.49 *** (1.20, 1.85) | 1.29 * (1.01, 1.64) |
| Do not know | 349 (22.3%) | 204 (31.1%) | 553 (24.9%) | 0.84 (0.67, 1.06) | 1.01 (0.77, 1.32) |
| Agree | 788 (50.3%) | 309 (47.2%) | 1097 (49.3%) | Ref |  |

\* p<0.05 \*\* p<0.01 \*\*\* p<0.001

Adults who completed the primary series vaccination but did not get a booster dose cited various reasons for their decisions. Approximately one-third (30.6%) mentioned that they had not received information for the next dose, while 30.5% said they did not need another dose. Less than a quarter of the participants (23.3%) reported other reasons, such as being sick due to COVID-19 or other illnesses, pregnancy, and fear of side effects from the booster (Figure 2).

## Discussion

This study is a community-based study highlighting contributing factors for COVID-19 booster vaccination uptake in Indonesia. We found that the coverage of the COVID-19 vaccination booster was 29.5%. This coverage is considerably low but comparable to reported national coverage, which was 29.6% during the study period (Kementrian Kesehatan RI, 2022a, 2022b). In addition, this study showed that hesitancy for a booster dosage is universal across provinces. Still, unwillingness is lower outside of Java Island. Studies have shown that those who lived in rural areas were less likely to get booster dosage due to difficult access to services that revealed inequality within the countries (Bennett et al., 2022; Saelee et al., 2022).

Nevertheless, these disparities are also seen in the United States for general COVID-19 vaccination (Gaffney et al., 2022). Thus, targeted strategies are crucial to address gaps in the coverage between Java and outside Java, also between rural and urban communities. This study found that individuals who perceived an upcoming new wave of COVID-19, disagreed with the end of the pandemic, were employed in the formal sector, had experienced side effects from previous vaccinations, and had a history of COVID-19 were more likely to receive a booster dose.

A previous study reported that essential workers such as teachers, healthcare professionals, and police had higher booster vaccination coverage compared to the non-essential ones (Lu et al., 2023), and the mandate for vaccination in the workplace resulted in increasing booster vaccination uptake (Bennett et al., 2022). However, other studies reported that having a history of COVID-19 (Lu et al., 2023) and having a side effect from previous vaccination were associated with lower uptake of booster vaccination, which contradicts our finding. In our study, people who take the booster dose dominated with higher education and work in the formal sector, which might have positive attitude on vaccination, trust on vaccine, have low concern on side effect and worry about the Pandemic as it explained by Geers (Geers et al., 2022) which found that experiencing side effect of primary vaccine is uncorrelated with COVID-19 booster intention They might have better access to information on COVID-19 and services related to booster dosage as also found in other studies (Abouzid et al., 2022; Thomas and Darling, 2021). This influences their perception towards a new wave and the end of the pandemic. It also affects their intention to keep the booster dose even though they had experienced COVID-19 (Jairoun et al., 2022) and the side effects of previous vaccination and belief in vaccination (Bennett et al., 2022; Jairoun et al., 2022). In addition, our result showed that low education emerged not to get booster vaccinations which, as reported by Thomas and Darling, this group were less trusting of the vaccine (Thomas and Darling, 2021). This finding suggests that barriers to access might remain, and efforts tailored to reach these groups were needed to increase uptake.

Our analysis did not find an association between infection risk, existing comorbidity, age, gender, and the booster dosage shot, which were inconsistent with previous findings on general COVID-19 vaccine acceptance determinants (Moola et al., 2021). While other studies reported that females were likely not vaccinated (Abouzid et al., 2022; Bennett et al., 2022; Lazarus et al., 2020), our findings showed no disparities in gender and age for booster dose uptake. This lack of disparity may be attributed to the national effort toward gender equality resulting in considerable progress in literacy, school enrollment, and employment (Afkar et al., 2020). Our result may also suggest that persons who complete the primary series vaccination, both for those with or without comorbidity, believe they have already been protected and do not need a booster (Abouzid et al., 2022; Lu et al., 2023). Furthermore, only 33.5% of participants perceived themselves as at risk, and only 12.4% expressed fear of transmission. Thus, it is crucial to encourage people, particularly those with comorbidity, to get a booster dose.

While this study provides a snapshot of booster vaccination status and perception at a point in time, it is important to note that both may change over time due to extensive campaigns and enforcement of regulations to increase vaccination uptake. It is worth noting that 16.3% of booster vaccination data were self-reported, which may be subject to biases and misreporting. The cross-sectional design limits our analysis to establish a temporal relationship.

Despite these limitations, this research contributes to our understanding that the current booster dose coverage in four provinces was not optimal, and disparities in geographic and other factors are apparent in booster dose uptake. Moreover, the perception was that the pandemic had ended and that no upcoming new wave had hindered the uptake of booster doses, leading to complacency regarding the low threat of COVID-19 (Wirawan et al., 2022).

It is important to note that the primary function of the booster dosage was not to protect against SARS-Cov2 infection but mainly to prevent disease severity, hospitalization, and fatality (Antonelli et al., 2022; Ioannou et al., 2022; Lin et al., 2023, 2022; Thompson, 2022; Tseng et al., 2022. Studies have shown that booster vaccination has the highest benefit impact on the elderly and high-risk groups with comorbidities and immunocompromised conditions (Johnson et al., 2022; Kelly et al., 2022). Therefore accelerating the booster dose coverage to those with the highest risk for severe disease should be considered (García-Botella et al., 2022). Shifting booster vaccination program from population-wide into a targeted vaccination strategy, which includes high-risk groups of comorbidities, immunocompromised people, and older age (Beaney et al., 2022; Dessie and Zewotir, 2021; Grasselli et al., 2020; Johnson et al., 2022), may yield the higher impact on reducing disease severity, hospitalization, and morbidity.

However, as our study found that the coverage of booster doses was lowest among the elderly age group, booster vaccination campaigns need to be tailored to this targeted population to reach the vaccine uptake determinant (Moola et al., 2021). Additionally, we need to promote community-based and faith-organizations collaboration in the national, provincial, and local health departments to put in place many initiatives that have been proven to increase the coverage of booster doses (Reitsma et al., 2021; Ryerson et al., 2021), This is on top of the previously proven strategies used to catch up the primary vaccination series. Moreover, there is a need for evidence of the cost-effectiveness of vaccine boosters between the general population and high-risk groups before continuing the administration of population-wide booster doses. A population-wide strategy may result in a marginal impact compared to the investment cost (Bardosh et al., 2022).

## Data Availability

All data produced in the present study are available upon reasonable request to the authors

## Acknowledgement

Directorate of Surveillance and Health Quarantine at the Ministry of Health, Republic of Indonesia, Provincial and District Health Offices in West Java, Banten, East Java and South Sulawesi Province for supporting the research. The Centre of Policy and Health Management, Dr Hanevi Jasri and dr. Muhammad Hardhantyo who contributed in facilitating the process.

## Disclaimer

The findings and conclusions in this report are those of the author(s) and do not necessarily represent the official position of the U.S. Centers for Disease Control and Prevention.

## Data availability

All data produced in the present study are available upon reasonable request to the corresponding authors

